# Intention and preference for long-acting injectable PrEP among MSM in the Netherlands: a diffusion of innovation approach

**DOI:** 10.1101/2022.11.11.22282218

**Authors:** Haoyi Wang, Hanne M. L. Zimmermann, David van de Vijver, Kai J. Jonas

**Affiliations:** Department of Work and Social Psychology, Maastricht University, Maastricht, The Netherlands; Viroscience Department, Erasmus Medical Centre, Rotterdam, The Netherlands; Department of Infectious Diseases, Public Health Service of Amsterdam, Amsterdam, The Netherlands

**Keywords:** MSM, Long-acting injectable PrEP, Diffusion of Innovation, suboptimal PrEP adherence, inte

## Abstract

Long-acting injectable PrEP (LAI-PrEP) is efficacious in preventing HIV among MSM and will be soon available in Europe. This study investigates the intention to use LAI-PrEP among MSM in the Netherlands by employing a diffusion of innovation approach, comparing PrEP naïve, discontinued MSM, oral PrEP early adopters and majority users. 309 MSM were surveyed on their intention and preference of LAI-PrEP. 83% showed high/very-high interest of, and 63% showed high/very-high intention to use LAI-PrEP. Early-adopters of oral PrEP use did not show increased intention to use LAI-PrEP and neither did PrEP naïve nor PrEP discontinued MSM, indicating a missing innovator effect for LAI-PrEP. Among the 218 current oral PrEP users, suboptimal oral PrEP adherence determined LAI-PrEP preference but did not determine LAI-PrEP intention. To reach the full potential of LAI-PrEP, a targeted strategy towards current oral PrEP users with suboptimal adherence seems indicated.

## Introduction

In the Netherlands, the HIV epidemic among MSM population is declining (1), and the oral HIV pre-exposure prophylaxis (PrEP) has been structurally introduced and included in the national oral PrEP project via the Public Health Services institutions in 2019 (1, 2). Recently in 2022, Public Health Services institutions in the Netherlands have estimated that there are around 30,000 MSM living in the Netherlands who meet the Dutch eligibility criteria of oral PrEP use (3). Yet, the available 8,500 slots of oral PrEP via the Public Health Services are already more or less taken (4). Among these current PrEP using MSM, most of them started to use oral PrEP through this structural delivery (hereinafter “majority oral PrEP users”). Yet, among this group there is a relevant proportion who had already used oral PrEP before the structural introduction in 2019 (hereinafter “the early adopters”) via different routes such as the Amsterdam PrEP (AMPrEP) demonstration project, informal individual procurement or via activist groups (4-7). Despite the increasing efforts of PrEP delivery, there are still MSM who had never used PrEP (hereinafter “PrEP naïve”) although they do meet the Dutch PrEP eligibility criteria, such as those who were not interested daily medication uptake (8, 9). Finally, there is a small, but relevant group of MSM who discontinued oral PrEP use for various reasons but who can still be considered to be at risk (10-13).

In addition, despite oral PrEP has proven a high efficacy in both daily use and on-demand use (14), its effectiveness decreases with a lower adherence (15). A recent study by Jourdain et al. has reported the real-world effectiveness of oral PrEP with an overall effectiveness of 60%, and differs by the user adherence from France, to range from 18% (low adherence) to 93% (high adherence) (16). Unfortunately, suboptimal adherence to oral PrEP is common among MSM (11, 16, 17). In the Netherlands, two of the main reasons behind suboptimal oral PrEP adherence are the low willingness to take medication daily required for oral daily PrEP (18) or forgetting to take it when taking PrEP on-demand (11, 19). As a consequence, MSM who are on oral PrEP can still get infected with HIV, especially among those with suboptimal adherence of oral PrEP. Taken together, there is still potential to further reduce HIV infections among MSM by means of biomedical prevention approaches.

Recently, bi-monthly injecting Cabotegravir as a long-acting injectable PrEP (LAI-PrEP), which has shown a superior effectiveness against HIV compared to oral PrEP with a 66% additional reduction in incident HIV infections (20), was authorised by the Food and Drug Agency in United States in 2021 and was just authorised by European Medicines Agency in Europe (21, 22). Therefore, LAI-PrEP may further reduce the current HIV epidemic among MSM by alleviating adherence problems of oral PrEP (20, 23). Researchers also hypothesised that LAI-PrEP can motivate PrEP naïve MSM who did not use oral PrEP or MSM who discontinued using oral PrEP due to the lack of daily medication uptake motivation to start using PrEP. As a result, additional HIV infections among PrEP naïve and PrEP discontinued MSM can be prevented (21, 23).

To determine at risk populations that can be served by LAI-PrEP better, a diffusion of innovation (DOI) theory can be helpful. DOI, seeks to understand how new ideas, practices and technologies spread through social systems and become accepted or normative (24). In the past, we observed an innovator effect from the early adopter stage to the majority stage for the oral PrEP uptake in the Netherlands: There were MSM who had used oral PrEP as the early adopters, and this had led to a major increase in PrEP uptake prior to the formal availability of PrEP in the Netherlands (4, 5, 8, 25). It is thus indicated to investigate if the same innovator effect would be observed again for the LAI-PrEP. More specifically, it needs to be assessed if those early adopters of oral PrEP (from more or less 6 years ago) would be again early adopters of LAI-PrEP in the near future.

This study thus sought to employ the DOI approach to investigate the determinants of the intention of LAI-PrEP among a cohort of MSM in the Netherlands by focusing on the comparisons among MSM who are PrEP-naïve, discontinued oral PrEP, early adopters, and the majority oral PrEP users. In addition, this study aimed to investigate, together with other determinants of LAI-PrEP use intention, how current oral PrEP adherence impacts on oral PrEP users’ intention and preference for using LAI-PrEP when it became officially available in the Netherlands.

## Methods

### Participants and Procedures

This study has a cross-sectional study design nested within a cohort study. All MSM living in the Netherlands in this study were recruited from an ongoing cohort which was established in 2017 (T0) and received follow-up questionnaires at three (T1) and six months (T2) to understand oral PrEP use among MSM (8, 9), and in 2020 during the COVID-19 pandemic (T3) to understand their sexual behaviours and experiences with PrEP during the pandemic. Participants were contacted again in June 2022 to complete an online survey on their awareness, interest, intention, and preference to use LAI-PrEP (T4). Only MSM without a positive HIV status and currently living in the Netherlands were included in this study. Participants were given the option of entering a raffle to win a €20 gift card. This study was approved by the Ethics Review Committee Psychology and Neuroscience of Maastricht University (ERCPN-174_10_12_2016). Informed consent was provided by all participants.

### Measures

#### LAI-PrEP awareness, interest, intention, preference, mixed-use intention, and tourism acceptance

Participants were asked whether they are aware of LAI-PrEP (“Yes”/ “No”). After providing the answer regardless of their LAI-PrEP awareness, all participants were provided a detailed introduction on 1) what is LAI-PrEP, 2) how to take LAI-PrEP, and 3) the differences between LAI-PrEP and oral PrEP including mechanisms, effective components, effectiveness in HIV prevention, and potential side effects.

Then, participants were asked whether they are interested in using LAI-PrEP when it becomes officially available with the national health care system (“hereinafter LAI-PrEP interest), measured on a 1-5 Likert scale (with 1 = “definitely not” and 5 = “definitely yes”). We also dichotomised LAI-PrEP interest as “Yes” and “No and not sure”.

Participants were further asked how likely is it that they will start using LAI-PrEP if it becomes officially available (hereinafter LAI-PrEP intention), measured on a 1-5 Likert scale (with 1 = “definitely not” and 5 = “definitely yes”). We also categorised LAI-PrEP intention as “High/very high”, “Not sure” and “Low/very low”.

In addition, participants were asked which regimen option they will prefer to use if they are all available (hereinafter LAI-PrEP preference), given the options of “LAI-PrEP”, “Daily oral PrEP”, “On demand oral PrEP”, “Mixed use of LAI-PrEP and oral PrEP”, and “Do not want to use PrEP at all”.

Participants who showed “High/Very high” or “Not sure” LAI-PrEP interest were asked “Imagine LA PrEP is available more easily and cheaper in other countries compared to where you live now, would you try to obtain LAI-PrEP there” (hereinafter LAI-PrEP tourism acceptance), and was measured and dichotomised as “Yes” and “No”. They were also asked their intention to use LAI-PrEP after given a scenario of “Due to the concerns of the potential drug resistance after stopping with LAI-PrEP, you may be asked to take oral PrEP daily for 180 days after you decide to no longer take the LAI-PrEP”, given the pharmacokinetic tail phase of LAI-PrEP (26, 27) (hereinafter LAI-PrEP intention with 180 days obligation), and was measured as same as the LAI-PrEP intention.

Lastly, participants who indicated “High/Very high” or “Not sure” LAI-PrEP intention were asked if they will use LAI-PrEP and oral PrEP together (hereinafter mixed use of LAI-PrEP), given the options of “No, I would like to only use LA PrEP”, “Yes, I would like to use both oral PrEP and LAI-PrEP at the same time”, “Yes, I would like to use both LAI-PrEP and use oral PrEP but at different time periods in a year”, and “I am not sure”.

#### Demographic variables

Among demographic variables, age was measured in years and was dichotomised as “< 40-year-old” and “>= 40-year-old”. Sexual orientation was measured and categorised as “Gay”, “Bisexual” and “Other”. For education, we defined MSM who did not have a high-school diploma as “Low”, and who had high school and higher than high-school education as “High”, following the definition of education level in the Netherlands (28). Financial status was measured and dichotomised as “Low” and “Living comfortable/very comfortable with current income”. Participants were also asked if they were born in the Netherlands and provide their migrant status as “Dutch native”, “European” and “Non-European”. Residence location was measured and dichotomised as “Main urban cities” and “The rest of the country” based on the provided 4-digit postal codes.

#### Behavioural variables

Relationship status was measured and categorised as “Single”, “Having dates”, “In a monogamous relationship” and “In an open/polyamorous relationship”. Having a steady partner with diagnosed HIV was measured as “Yes” and “No”. Having a non-steady partner with diagnosed HIV was measured as “Yes”, “No” and “Not sure”. Ever having any type of sexually transmitted infections (STIs) was measured as “Yes, within the previous 12 months”, “Yes, more than 12 months” and “Never”. Ever engaged in chemsex was measured as “Yes” and “No”. We divided ever engaged in transactional sex into 1) ever paid for sex and 2) ever got paid for sex” and measured these two variables as “Yes, within the previous 12 months”, “Yes, more than 12 months” and “Never”.

In addition, following the DOI approach, based on our longitudinal data, we measured the current oral PrEP use status together with the initiation of oral PrEP uptake. We categorised oral PrEP use status as “Early adopter” (based on an initiation prior to the structural availability in 2019 in the Netherlands), “Majority oral PrEP user” (initiation after 2019), “PrEP naïve” and “PrEP discontinued” (any PrEP use discontinued).

Oral PrEP adherence was also assessed among the current oral PrEP users their oral PrEP use modes: daily, on-demand, and Tuesday-Thursday-Saturday-Sunday (TTSS) scheme. For daily and TTSS scheme oral PrEP users, we measured their adherence by asking “How often do you miss a pill”; and defined “Never” and “Miss using pill once or twice a month” as “Good adherence”. For on-demand oral PrEP users, we assessed their PrEP adherence by asking 1) “How often do you just take 1 pill in advance”, 2) “How often did you have sex less than 2hrs after the first 2 pills”, and 3) “How often do you forget to take a pill 24 and/or 48hrs after”. We defined on-demand oral PrEP users who “Always” and “Almost all the time” take 1 pill in advance and have sex less than 2hrs after the first 2 pills, and who “Never” and “Occasionally” forget to take a pill 24 and/or 48hrs after with “Good adherence”. An overall oral PrEP adherence was re-coded and dichotomised as “Good adherence” and “Suboptimal adherence”.

#### Psycho-social variables

We applied the Big Five personality inventory scale (29) to measure associations between the five main personalities (Extraversion, Agreeableness, Conscientiousness, Neuroticism and Openness) and our endpoints. All personality assessment items were measured on a 5-point Likert scale (with 1 = “Disagree strongly” and 5 “Agree strongly”).

### Statistical analysis

We first generated two descriptive analyses for all included variables for 1) all participants and 2) by their LAI-PrEP intention, which was dichotomised as “High/very high” and the rest of scales. We then generated another two descriptive analyses for 3) the current oral PrEP users and 4) by their LAI-PrEP intention status. Fisher’s exact test and chi-square test were used to test the differences of the included variables between different LAI-PrEP intention status.

We conducted three multivariable logistic regression analyses to identify demographic, behavioural and psycho-social determinants of 1) LAI-PrEP intention among MSM (total included samples), 2) LAI-PrEP intention among current oral PrEP users, and 3) LAI-PrEP preference among current oral PrEP users. Given the relatively small proportion of the participants showing “not sure” and “low/very low” intention for LAI-PrEP, to better inform public health measures and actions, we dichotomised LAI-PrEP intention as “High/very high” and the rest of the scales for analyses with LAI-PrEP intention as the endpoint. For LAI-PrEP preference as the endpoint, following the same approach, we also dichotomised it as “LAI-PrEP or mixed use of LAI-PrEP and oral PrEP” vs. “Oral PrEP”, given no current oral PrEP user was found to not want to use PrEP.

For each of these regression analyses, we first conducted an univariable logistic model with each of the selected demographic, behavioural and psycho-social determinants to investigate potential correlation with each endpoint. Next, we retained all variables with p<0.10 in a multivariable full model (30). Then, a manual stepwise backward selection approach was applied to select the explanatory variables in a multivariable final model. We considered all variables with p<0.05 statistically significant. For the current oral PrEP user analyses, current oral PrEP use status and current oral PrEP adherence were retained regardless of the significance, given they were the key variables of interest in this study, and were considered relevant for public health with p<0.10. All analyses were conducted in R (version R 4.2.1).

## Results

### Study characteristics

All 766 cohort participants from T0 were invited to participate in this study, of which 327 of them completed this survey. We excluded 18 participants from this study due to 1) having received an HIV diagnosis (n=6) and not currently living in the Netherlands (n=12), resulting in 309 participants in total included in this study. The flowchart of data collection at each study point was described in Figure 1.

**Figure 1.**
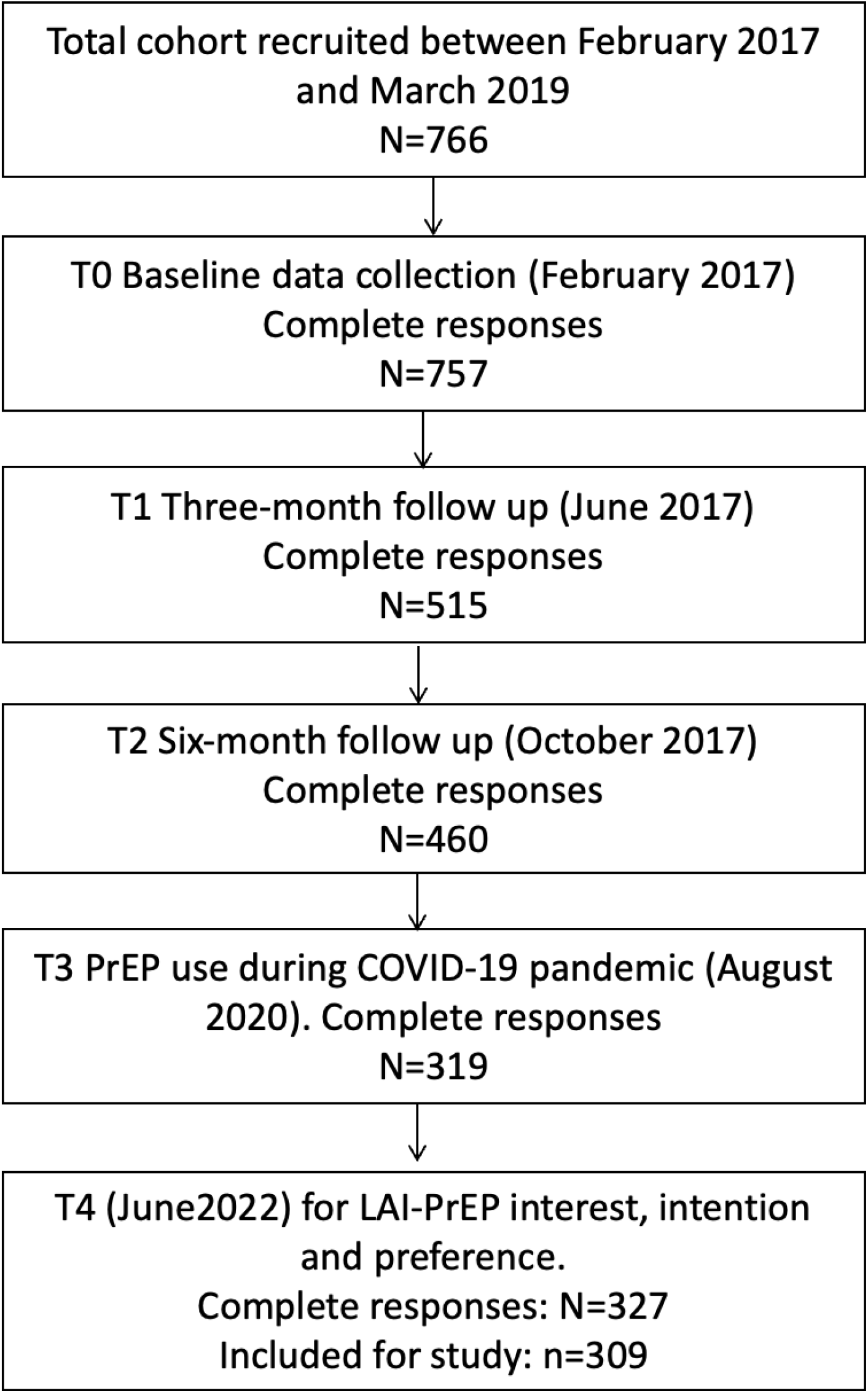
Flowchart of participants per data point. Note: LAI-PrEP = long-acting injectable pre-exposure prophylaxis. Detailed information for each data point see our previous study (9).

Of the 309 MSM included, 31% (95 of 309) were <40-year-old (median age=47, interquartile range 38-54), 15% (45 of 309) were PrEP-naïve, 15% (46 of 309) discontinued oral PrEP, and 45% (128 of 309) were early adopters. Among them, MSM with different age groups (p=0.002), who with different immigration backgrounds (p=0.04), who had different oral PrEP use statuses (p=0.03), who had different STIs diagnoses (p=0.04), and who had/had not engaged in chemsex (p=0.02) had significantly different LAI-PrEP intention. More participants’ characteristics by LAI-PrEP intention can be found in Table 1.

**Table 1.** Study characteristics of all included MSM.

| Variables | Total sample<br>(n=309) |  | High or very high intention<br>to use LAI-PrEP (n=194) |  | Not sure, Low or very<br>low intention to use LAI-PrEP (n=115) |  | P-value<br>(Fisher's exact test<br>or chi-square test) |
| --- | --- | --- | --- | --- | --- | --- | --- |
|  | n/medi<br>an | %/IQR | n/median | %/IQR | n/median | %/IQR |  |
| Age |  |  |  | § |  |  |  |
| <40-year-old | 95 | 30.7 | 72 | 37,1 | 23 | 20,0 | 0.002 |
| >=40-year-old | 214 | 69.3 | 122 | 62,9 | 92 | 80,0 |  |
| Sexual orientation |  |  |  |  |  |  |  |
| Gay | 287 | 93.2 | 181 | 93,3 | 106 | 92,2 | 0.67 |
| Bisexual | 13 | 4.2 | 9 | 4,6 | 4 | 3,5 |  |
| Other | 9 | 2.6 | 4 | 2,1 | 5 | 4,3 |  |
| Education |  |  |  |  |  |  |  |
| Low | 33 | 10.7 | 23 | 11,9 | 10 | 8,7 | 0.45 |
| High | 276 | 89.3 | 171 | 88,1 | 105 | 91,3 |  |
| Employment |  |  |  |  |  |  |  |
| Employed | 279 | 90.3 | 178 | 91,8 | 101 | 87,8 | 0.53 |
| Unemployed | 5 | 1.6 | 3 | 1,5 | 2 | 1,7 |  |
| Retired | 19 | 6.1 | 9 | 4,6 | 10 | 8,7 |  |
| Student | 6 | 1.9 | 4 | 2,1 | 2 | 1,7 |  |
| Finance |  |  |  |  |  |  |  |
| Low | 93 | 30.1 | 64 | 33,0 | 29 | 25,2 | 0.16 |
| Living comfortable/very<br>comfortable with current income | 216 | 69.9 | 130 | 67,0 | 86 | 74,8 |  |
| Migrant status |  |  |  |  |  |  |  |
| Dutch | 249 | 80.6 | 148 | 76,3 | 101 | 87,8 | 0.04 |
| European | 20 | 6.5 | 16 | 8,2 | 4 | 3,5 |  |
| Non-European | 40 | 12.9 | 30 | 15,5 | 10 | 8,7 |  |
| Geo-location |  |  |  |  |  |  |  |
| Main urban cities | 211 | 68.3 | 128 | 66,0 | 83 | 72,2 | 0.31 |
| The rest of the country | 98 | 31.7 | 66 | 34,0 | 32 | 27,8 |  |
| Oral PrEP use status |  |  |  |  |  |  |  |
| Early adopter | 128 | 41.4 | 86 | 44,3 | 42 | 36,5 | 0.03 |
| Majority oral PrEP user | 90 | 29.1 | 62 | 32,0 | 28 | 24,3 |  |
| PrEP naïve | 45 | 14.6 | 21 | 10,8 | 24 | 20,9 |  |
| PrEP discontinued | 46 | 14.9 | 25 | 12,9 | 21 | 18,3 |  |
| LAI-PrEP awareness |  |  |  |  |  |  |  |
| Yes | 124 | 40.1 | 82 | 42,3 | 42 | 36,5 | 0.16 |
| No | 185 | 59.9 | 112 | 57,7 | 73 | 63,5 |  |
| LAI-PrEP Interest |  |  |  |  |  |  |  |
| Yes | 256 | 82.8 | 192 | 99,0 | 64 | 55,7 | <0.001 |
| No, not sure | 53 | 17.2 | 2 | 1,0 | 51 | 44,3 |  |
| LAI-PrEP Intention |  |  |  |  |  |  |  |
| Yes | 194 | 62.8 | NA | NA | NA | NA | NA |
| No | 76 | 24.6 | NA | NA | NA | NA |  |
| Not sure | 39 | 12.6 | NA | NA | NA | NA |  |
| PrEP regimens preference |  |  |  |  |  |  |  |
| Do not want to use PrEP at all | 13 | 4.2 | 2 | 1,0 | 11 | 9,6 | <0.01 |
| LAI-PrEP | 152 | 49.2 | 132 | 68,0 | 20 | 17,4 |  |
| Daily oral PrEP | 37 | 12.0 | 12 | 6,2 | 25 | 21,7 |  |
| On-demand oral PrEP | 59 | 19.1 | 15 | 7,7 | 44 | 38,3 |  |
| Mixed use of different regimens at different time | 48 | 15.5 | 33 | 17,0 | 11 | 9,6 |  |
| LAI-PrEP tourism acceptance** |  |  |  |  |  |  |  |
| Yes | 46 | 17.3 | 38 | 20,3 | 8 | 10,1 | 0.05 |
| No | 220 | 82.7 | 149 | 79,7 | 71 | 89,9 |  |
| Mixed use of LAI-PrEP* |  |  |  |  |  |  |  |
| Only LAI-PrEP | NA | NA | 121 | 54.8 | NA | NA | NA |
| Mixed use of LAI-PrEP and oral PrEP at different time | NA | NA | 31 | 13.9 | NA | NA |  |
| Mixed use of LAI-PrEP and oral PrEP at the same time | NA | NA | 3 | 1.4 | NA | NA |  |
| Not sure | NA | NA | 66 | 29.9 | NA | NA |  |
| LAI-PrEP intention with 180 days obligation** |  |  |  |  |  |  |  |
| Yes | 152 | 57.1 | 131 | 70,1 | 21 | 26,6 | <0.001 |
| No | 72 | 27.1 | 31 | 16,6 | 41 | 51,9 |  |
| Not sure | 42 | 15.8 | 25 | 13,4 | 17 | 21,5 |  |
| Relationship status |  |  |  |  |  |  |  |
| Single | 118 | 38.2 | 84 | 43,3 | 34 | 29,6 | 0.06 |
| Having dates | 12 | 3.9 | 9 | 4,6 | 3 | 2,6 |  |
| In a relationship | 78 | 25.2 | 43 | 22,2 | 35 | 30,4 |  |
| In an open/polyamorous relationship | 101 | 32.7 | 58 | 29,9 | 43 | 37,4 |  |
| Having steady partner with diagnosed HIV |  |  |  |  |  |  |  |
| Yes | 159 | 51.5 | 96 | 49,5 | 63 | 54,8 | 0.41 |
| No | 150 | 48.5 | 98 | 50,5 | 52 | 45,2 |  |
| Having non-steady partner with diagnosed HIV |  |  |  |  |  |  |  |
| Yes | 114 | 36.9 | 76 | 39,2 | 38 | 33,0 |  |
| No | 110 | 35.6 | 68 | 35,1 | 42 | 36,5 | 0.56 |
| Not sure | 85 | 27.5 | 50 | 25,8 | 35 | 30,4 |  |
| STIs diagnosis |  |  |  |  |  |  |  |
| Yes, within the previous 12 months | 102 | 33.0 | 74 | 38,1 | 28 | 24,3 | 0.04 |
| Yes, more than 12 months ago | 157 | 50.8 | 90 | 46,4 | 67 | 58,3 |  |
| Never | 50 | 16.2 | 30 | 15,5 | 20 | 17,4 |  |
| Ever engage in Chemsex |  |  |  |  |  |  |  |
| Yes | 132 | 42.7 | 93 | 47,9 | 39 | 33,9 | 0.02 |
| No | 177 | 57.3 | 101 | 52,1 | 76 | 66,1 |  |
| Buy sex |  |  |  |  |  |  |  |
| Yes, within the previous 12 months | 34 | 11.0 | 18 | 9,3 | 16 | 13,9 | 0.32 |
| Yes, more than 12 months ago | 16 | 5.2 | 12 | 6,2 | 4 | 3,5 |  |
| Never | 259 | 83.8 | 164 | 84,5 | 95 | 82,6 |  |
| Paid sex (n=305) |  |  |  |  |  |  |  |
| Yes, within the previous 12 months | 31 | 10.0 | 22 | 11,3 | 9 | 7,8 | 0.14 |
| Yes, more than 12 months ago | 9 | 2.9 | 8 | 4,1 | 1 | 0,9 |  |
| Never | 256 | 85.8 | 161 | 83,0 | 104 | 90,4 |  |
| The Big Five personality inventory§ |  |  |  |  |  |  |  |
| Extraversion | 3.0 | [2.5-4.0] | 3.0 | [2.5-4.0] | 3.0 | [2.25-3.5] | 0.17 |
| Agreeableness | 4.0 | [3.5-4.5] | 4.0 | [3.5-4.0] | 4.0 | [3.5-4.5] | 0.94 |
| Conscientiousness | 3.5 | [3.0-4.5] | 3.5 | [3.0-4.5] | 3.75 | [3.0-4.5] | 0.90 |
| Neuroticism | 2.5 | [2.0-3.5] | 2.5 | [2.0-5.0] | 2.5 | [2.0-3.5] | 0.80 |
| Openness | 3.5 | [3.0-4.5] | 3.5 | [3.0-4.5] | 3.75 | [3.0-4.5] | 0.10 |

Note: \* indicates question was only asked for those who showed somehow LAI-PrEP intention. \*\* indicates question was only asked for those who showed somehow LAI-PrEP interest. § indicate variables were measured on a 1-5 agreeable scale (with 1 = “Disagree strongly” and 5 “Agree strongly”) and presented with median [IQR]. LAI-PrEP = long-acting pre-exposure prophylaxis. NA = not applicable.

Of the 218 current oral PrEP users, 27% (59 of 218) were <40-year-old, 59% (128 of 218) were early adopters and 10% (22 of 218) showed suboptimal current oral PrEP adherence. Among them, MSM with different age groups (p<0.001), who had different financial status (p=0.03), who had different immigration backgrounds (p=0.02), who had different STIs diagnoses (p=0.04), and who with more/less open personality characteristics had significant different LAI-PrEP intention. More current oral PrEP users’ characteristics by LAI-PrEP intention can be found in Table 2.

**Table 2.**
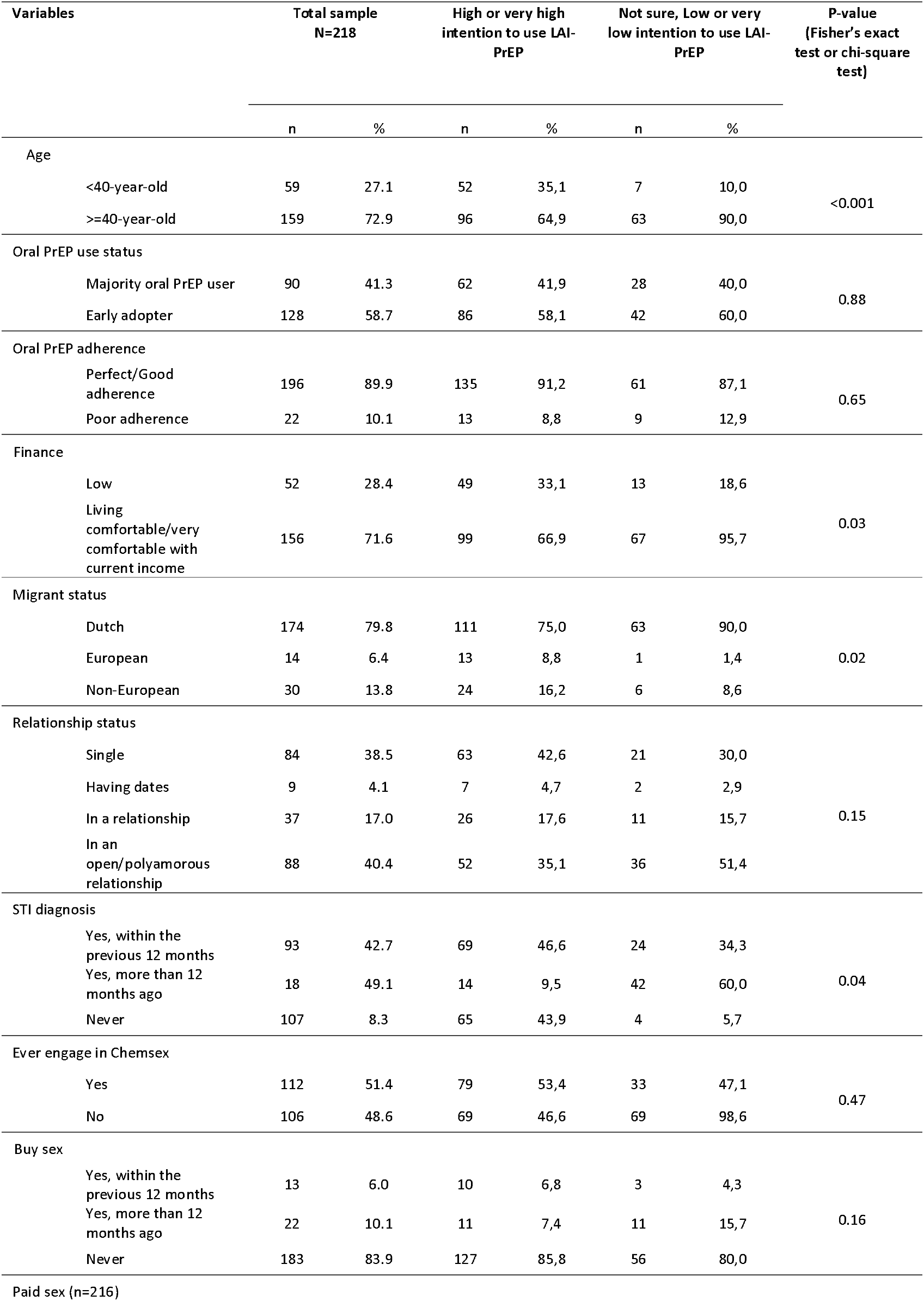
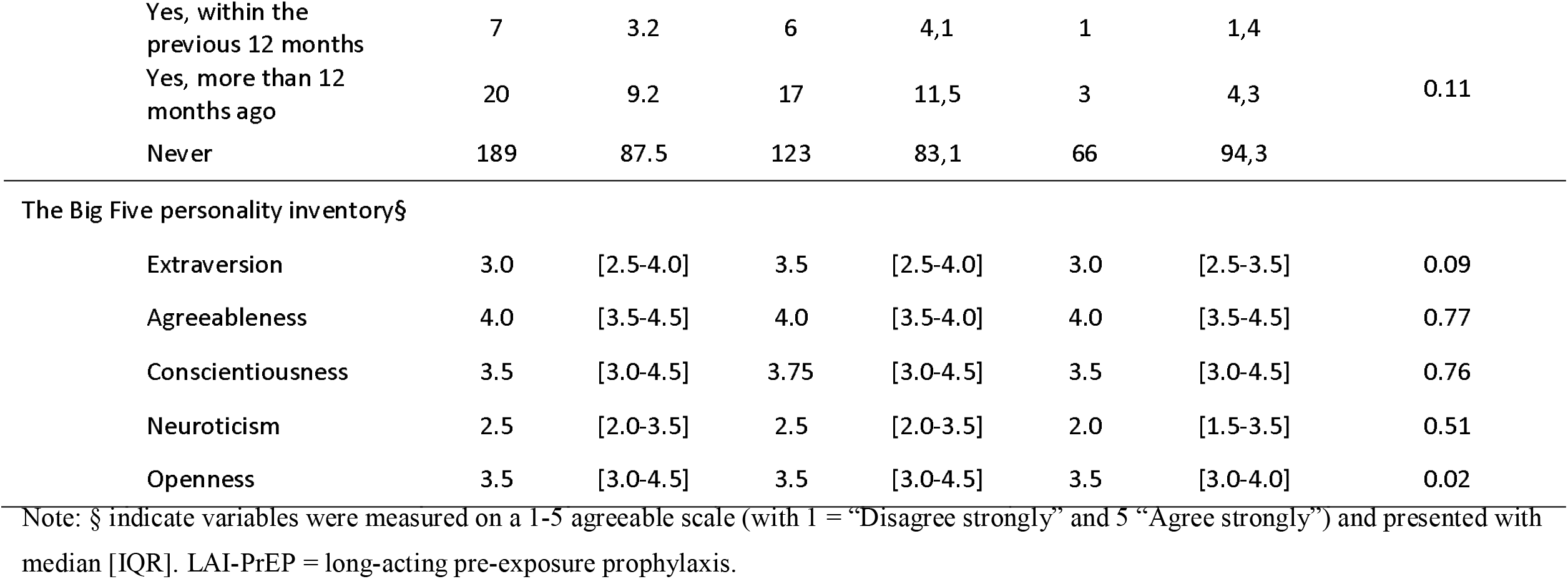
Study characteristics of MSM who were currently using PrEP.

### LAI-PrEP awareness, interest, intention, preference, mixed-use intention, and tourism acceptance

Among all the included MSM, 40% (124 of 309) were aware of LAI-PrEP, 83% (256 of 309) showed high/very high interest of, and 63% (194 of 309) showed high/very high intention to use LAI-PrEP, and 49% (152 of 309) would prefer to use LAI-PrEP, 16% (48 of 309) would prefer to mixed-use LAI-PrEP and oral PrEP. After being informed the potential 180-day obligation of using oral PrEP daily after discontinuing using LAI-PrEP, still 57% (152 of 309) reported high/very high intention to use LAI-PrEP. Only 17% (46 of 266) of those who indicated to be willing to travel to other countries for LAI-PrEP uptake. 14% (14 of 194) indicated would use both LAI-PrEP and oral PrEP at different periods in a year and only 1% indicated would use both LAI-PrEP and oral PrEP at the same time (3 of 194). For more information, see Table 1. Figure 2 summarises the frequency distribution of LAI-PrEP awareness, interest, and intention among MSM in general and by oral PrEP use status. Despite the overall moderate LAI-PrEP awareness and high LAI-PrEP interest and intention, early adopters of oral PrEP were found to have the highest LAI-PrEP awareness, interest, and intention, while PrEP naïve MSM were found to have the lowest LAI-PrEP awareness, interest, and intention.

**Figure 2.**
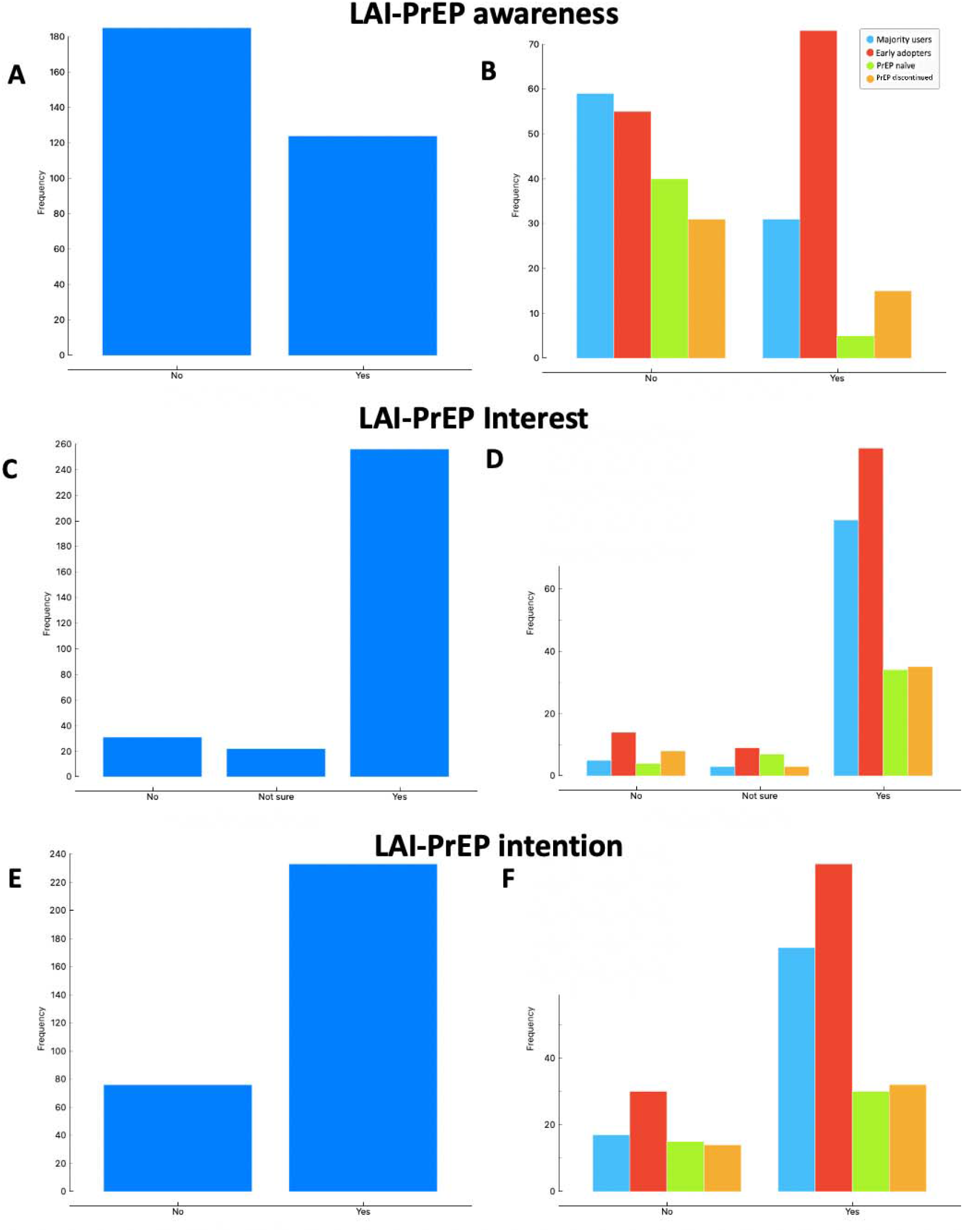
Distribution of (A) LAI-PrEP awareness among MSM, (B) LAI-PrEP awareness among MSM by oral PrEP use status, (C) LAI-PrEP interest among MSM, (D) LAI-PrEP interest among MSM by oral PrEP use status, (E) LAI-PrEP intention among MSM, and (F) LAI-PrEP intention among MSM by oral PrEP use status. Note: LAI-PrEP = long-acting injectable pre-exposure prophylaxis.

Among the current oral PrEP users, 48% (104 of 218) were aware of LAI-PrEP, 82% (105 of 218) showed high/very high interest of, and 67% (86 of 218) showed high/very high intention to use LAI-PrEP. 50% (110 of 218) would prefer to use LAI-PrEP and 18% (39 of 218) would prefer to use LAI-PrEP and oral PrEP at different time over the oral PrEP preference. For more information, see Table 2.

### Determinants of LAI-PrEP intention among MSM

For the LAI-PrEP intention endpoint, our final multivariable logistic regression analysis showed that MSM who were older than 40-year-old (adjusted odds ratio (aOR)=0.43, 95%CI=0.21 - 0.84), who were PrEP naïve (aOR=0.39, 0.16 - 0.92), who discontinued using PrEP (aOR=0.46, 0.18 - 1.14), and who never (aOR=0.03, 0.00 - 0.75) and who got paid for sex more than 12 months ago (aOR=0.05, 0.00 - 1.31) were less likely to have a “high” LAI-PrEP or “not sure” LAI-PrEP interest (aOR=0.01, 0.00 - 0.05). No personality characteristics were found to be statistically significant associated with the LAI-PrEP intention among all MSM included. For results obtained from univariable logistic regression analyses, see Table 3. Figure 3 showed the comparison of the intention to use LAI-PrEP by the current oral PrEP use status following the DOI approach through multivariable logistic modelling. Compared to the early oral PrEP adopters, only PrEP naïve MSM had a statistically lower LAI-PrEP intention, and no statistically significant difference was observed between early oral PrEP adopters, the majority oral PrEP users and PrEP discontinued MSM.

**Table 3.** Determinants of LAI-PrEP intention (high/very high vs. the rest of the scale) among MSM.

| Variables | Univariable model |  |  | Multivariable model |  |  |
| --- | --- | --- | --- | --- | --- | --- |
|  | OR | 95%CI | P-value | aOR | 95%CI | P-value |
| Age |  |  |  |  |  |  |
| <40-year-old | Ref |  |  | Ref |  |  |
| >=40-year-old | 0.42 | 0.24 - 0.71 | 0.002 | 0.43 | 0.21 - 0.84 | 0.01 |
| Sexual orientation |  |  |  |  |  |  |
| Gay | Ref |  |  |  |  |  |
| Bisexual | 1.32 | 0.42 - 4.96 | 0.65 |  |  |  |
| Other | 0.59 | 0.14 - 2.52 | 0.46 |  |  |  |
| Education |  |  |  |  |  |  |
| Low | Ref |  |  |  |  |  |
| High | 0.70 | 0.31 - 1.51 | 0.39 |  |  |  |
| Employment |  |  |  |  |  |  |
| Employed | Ref |  |  |  |  |  |
| Unemployed | 0.85 | 0.14 - 6.54 | 0.51 |  |  |  |
| Retired | 0.51 | 0.20,1.30 | 0.88 |  |  |  |
| Student | 1.53 | 0.22 - 8.29 | 0.86 |  |  |  |
| Finance |  |  |  |  |  |  |
| Low | Ref |  |  |  |  |  |
| Living comfortable/very comfortable with current income | 0.68 | 0.40 - 1.14 | 0.15 |  |  |  |
| Migrant status |  |  |  |  |  |  |
| Dutch | Ref |  |  |  |  |  |
| European | 1.00 | 0.969.74 | 0.08 |  |  |  |
| Non-European | 0.71 | 0.99 - 4.58 | 0.06 |  |  |  |
| Geo-location |  |  |  |  |  |  |
| Main urban cities | Ref |  |  |  |  |  |
| The rest of the country | 1.34 | 0.81 - 2.23 | 0.25 |  |  |  |
| Oral PrEP use status |  |  |  |  |  |  |
| Early adopter | Ref |  |  | Ref |  |  |
| Majority oral PrEP user | 1.08 | 0.61 - 1.49 | 0.79 | 0.70 | 0.34 - 1.42 | 0.33 |
| PrEP naïve | 0.42 | 0.21 - 0.85 | 0.01 | 0.39 | 0.16 - 0.92 | 0.03 |
| PrEP discontinued | 0.58 | 0.29 - 1.16 | 0.12 | 0.46 | 0.18 - 1.14 | 0.09 |
| Relationship status |  |  |  |  |  |  |
| Single | Ref |  |  |  |  |  |
| Having dates | 1.21 | 0.34 - 5.72 | 0.78 |  |  |  |
| In a relationship | 0.50 | 0.27 - 0.90 | 0.02 |  |  |  |
| In an open/polyamorous relationship | 0.55 | 0.31 - 0.95 | 0.03 |  |  |  |
| Having steady partner with diagnosed HIV |  |  |  |  |  |  |
| Yes | Ref |  |  |  |  |  |
| No | 1.24 | 0.78 - 1.97 | 0.36 |  |  |  |
| Having non-steady partner with diagnosed HIV |  |  |  |  |  |  |
| Yes | Ref |  |  |  |  |  |
| No | 0.81 | 0.47 - 1.40 | 0.45 |  |  |  |
| Not sure | 0.74 | 0.41 - 1.32 | 0.30 |  |  |  |
| STI diagnosis |  |  |  |  |  |  |
| Yes, within the previous 12 months | Ref |  |  |  |  |  |
| Yes, more than 12 months ago | 0.51 | 0.29 - 0.86 | 0.01 |  |  |  |
| Never | 0.57 | 0.28 - 1.16 | 0.12 |  |  |  |
| Ever engage in Chemsex |  |  |  |  |  |  |
| Yes | Ref |  |  |  |  |  |
| No | 0.56 | 0.34 - 0.89 | 0.02 |  |  |  |
| Buy sex |  |  |  |  |  |  |
| Yes, within the previous 12 months | Ref |  |  |  |  |  |
| Yes, more than 12 months ago | 0.38 | 0.09 - 1.32 | 0.14 |  |  |  |
| Never | 0.58 | 0.16 - 1.70 | 0.35 |  |  |  |
| Paid sex (n=305) |  |  |  |  |  |  |
| Yes, within the previous 12 months | Ref |  |  | Ref |  |  |
| Yes, more than 12 months ago | 0.31 | 0.02 - 2.04 | 0.30 | 0.05 | 0.00 - 1.31 | 0.07 |
| Never | 0.19 | 0.01 - 1.08 | 0.12 | 0.03 | 0.00 - 0.75 | 0.03 |
| The Big Five personality inventory§ |  |  |  |  |  |  |
| Extraversion | 1.26 | 0.97 - 1.62 | 0.08 |  |  |  |
| Agreeableness | 0.97 | 0.62 - 1.24 | 0.46 |  |  |  |
| Conscientiousness | 0.91 | 0.68 - 1.22 | 0.54 |  |  |  |
| Neuroticism | 1.04 | 0.82, 1.32 | 0.76 |  |  |  |
| Openness | 0.98 | 0.76 - 1.28 | 0.92 |  |  |  |

Note: § indicate variables were measured on a 1-5 agreeable scale (with 1 = "Disagree strongly" and 5 "Agree strongly"). aOR = adjusted odds ratio. CI = confident interval. OR = odds ratio.

**Figure 3.**
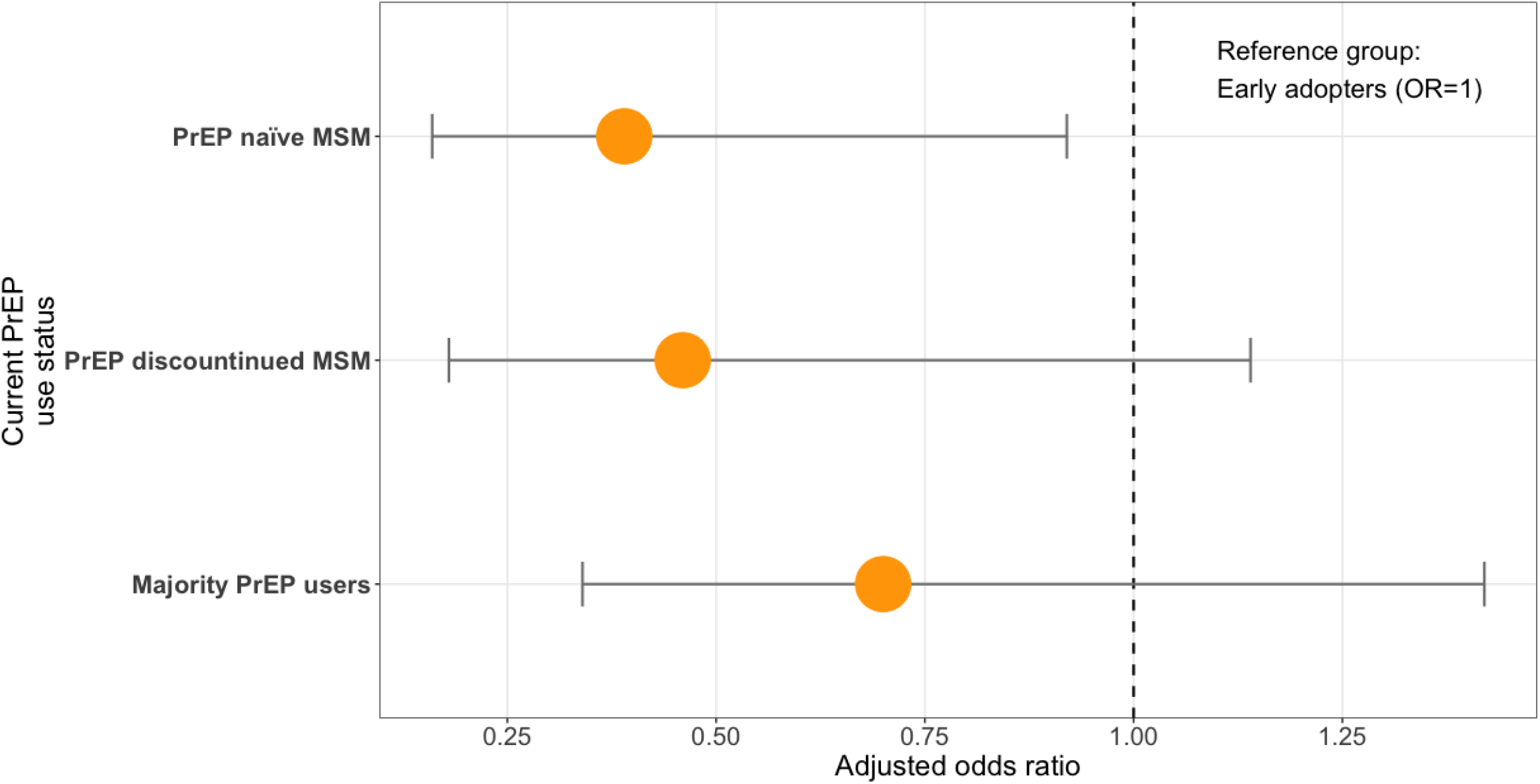
Intention to use long-acting injectable PrEP by current oral PrEP use status, following the confusion of innovation approach through multivariable logistic modelling. Note: PrEP = pre-exposure prophylaxis.

### Determinants of LAI-PrEP intention and preference among current oral PrEP users

For LAI-PrEP intention as endpoint among current oral PrEP users, our final multivariable logistic regression analysis showed that current oral PrEP users who were older than 40-year-old (aOR=0.25, 0.08 - 0.72), and who were living comfortably or very comfortably financially (aOR=0.91, 0.42 - 1.96) were less likely to have high LAI-PrEP intention. Being an early oral PrEP adopter (aOR=1.47, 0.72 - 3.02) and reporting suboptimal oral PrEP adherence (aOR=0.91, 0.42 - 1.96) were not associated with a high LAI-PrEP intention significantly. No personality characteristics were found statistically significant associated with the LAI-PrEP intention among the current oral PrEP users. For results obtained from univariable logistic regression analyses, see Table 4.

**Table 4.**
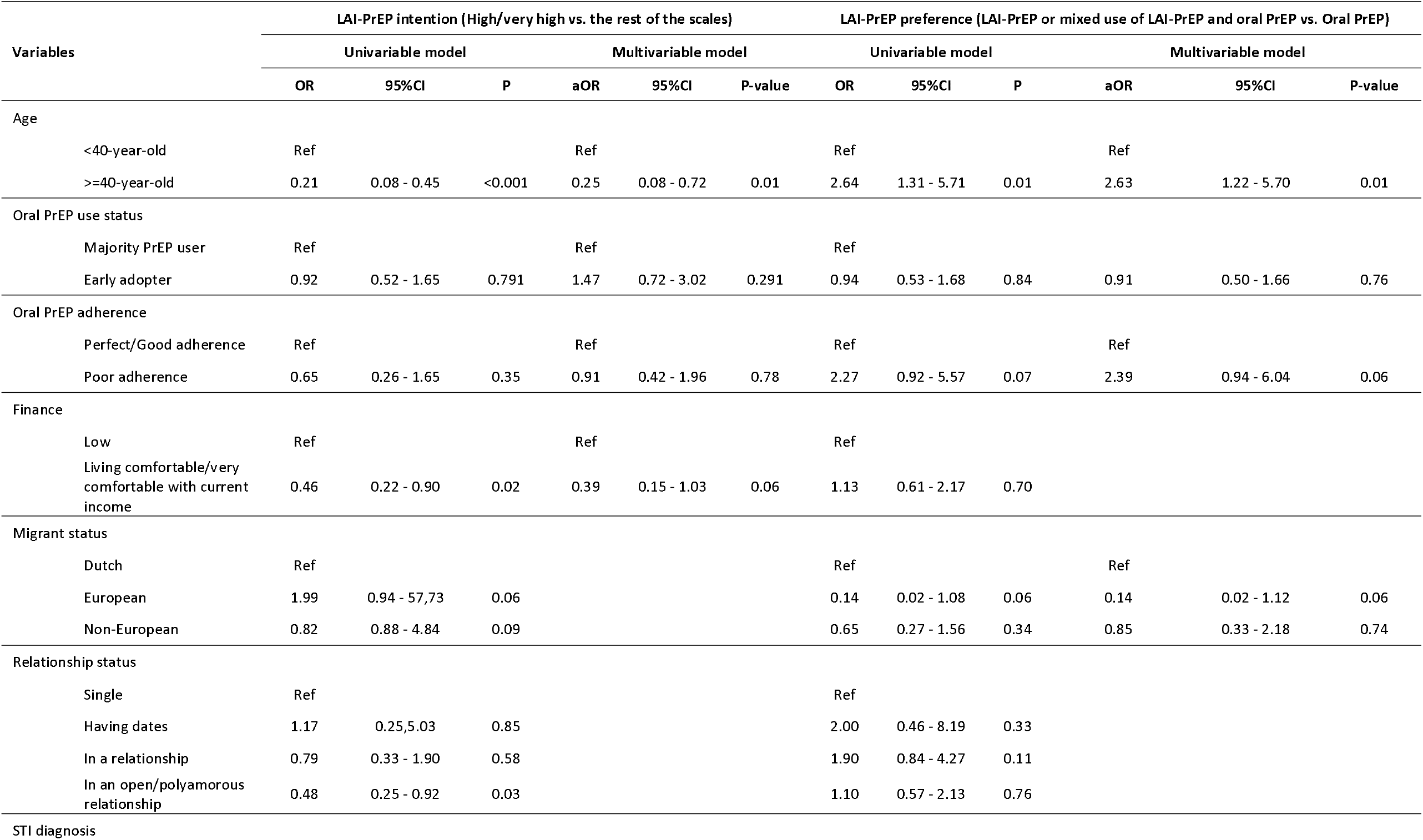
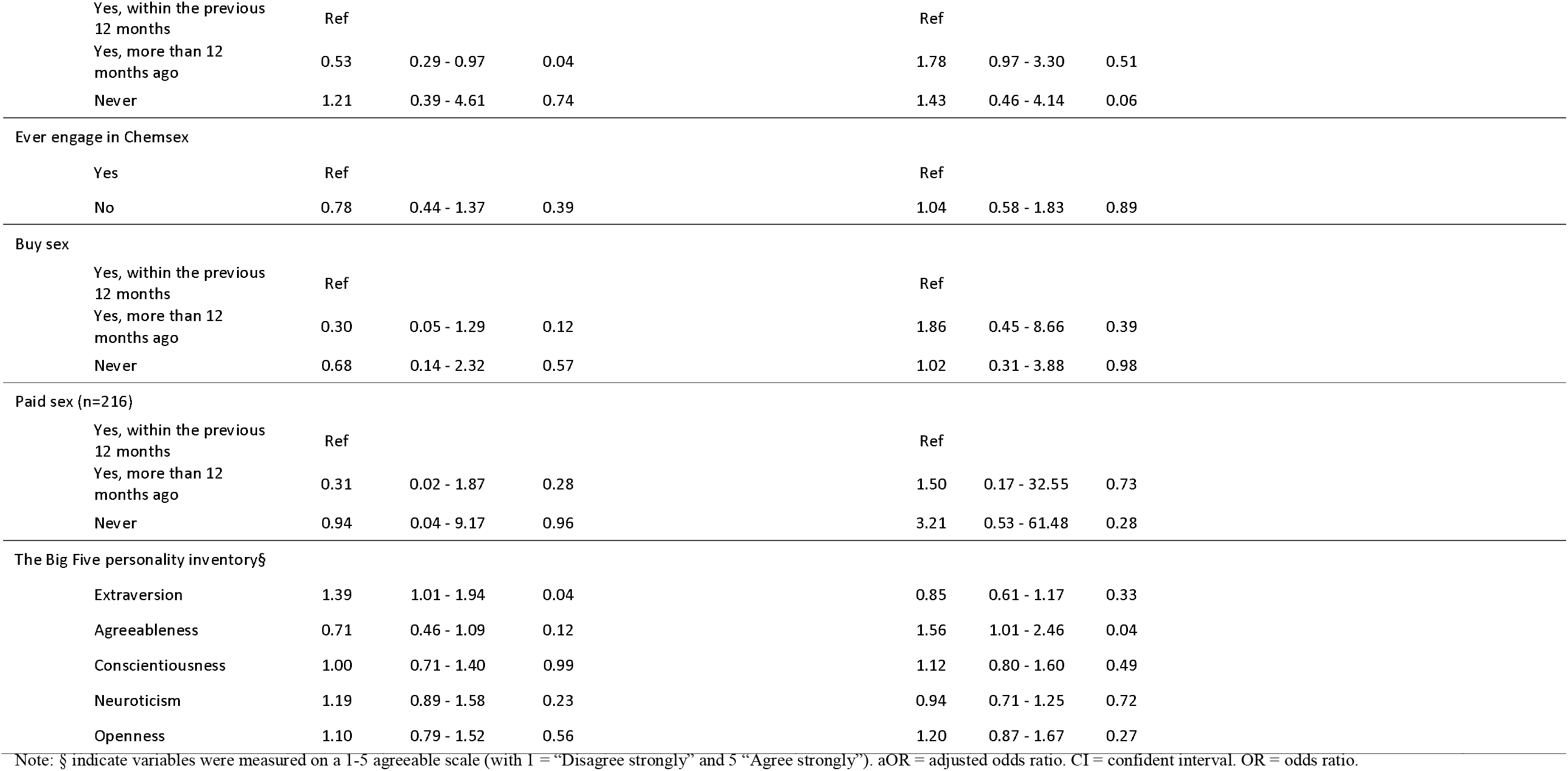
Determinants of LAI-PrEP intention and preference among current oral PrEP users.

For LAI-PrEP preference as endpoint among the current oral PrEP users, our final multivariable logistic regression analysis showed that current oral PrEP users who were older than 40-year-old (aOR=2.64, 1.12 - 5.70) were more likely to prefer to use LAI-PrEP. Current oral PrEP users who had a European migrant status (aOR=0.14, 0.02 - 1.12), on the other hand, were less likely to prefer to use LAI-PrEP. Similar to the LAI-PrEP intention, being an early adopter of oral PrEP was not predictive for a LAI-PrEP preference. What is noteworthy to mention is, for current oral PrEP users who had suboptimal oral PrEP adherence (aOR=2.39, 0.94 - 6.04, p=0.06) had the trend to be more likely to prefer using LAI-PrEP. No personality characteristics were found to be statistically significant associated with the LAI-PrEP preference among the current oral PrEP users. For results obtained from univariable logistic regression analyses, see Table 4. Figure 4 showed the comparison of the intention and preference to use LAI-PrEP by the current oral PrEP use status and current oral PrEP adherence through multivariable logistic modelling.

**Figure 4.**
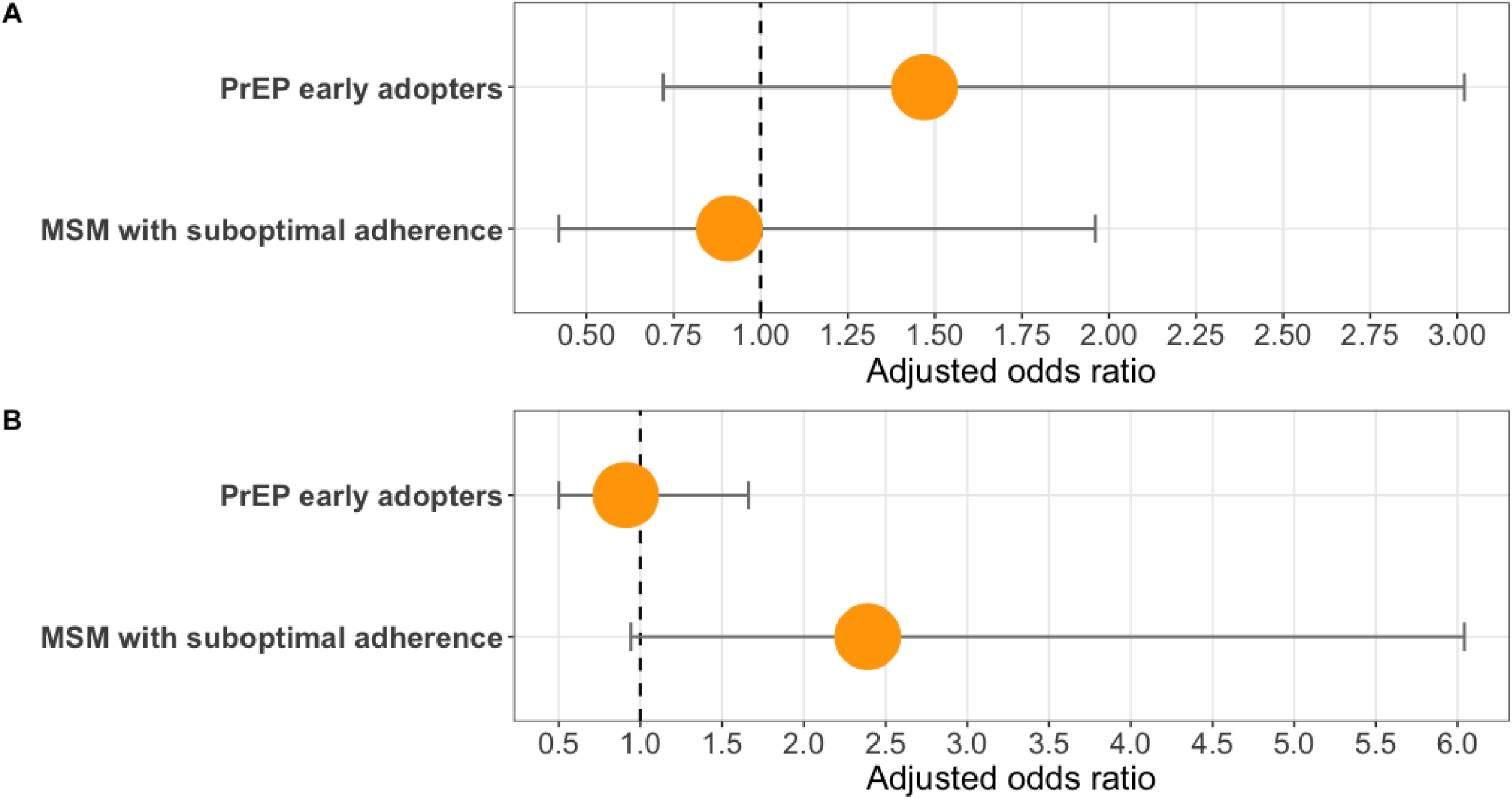
(A) Intention to and (B) preference of using long-acting injectable PrEP by current oral PrEP use status, following the confusion of innovation approach, and current oral PrEP adherence through multivariable logistic modelling. Note: PrEP = pre-exposure prophylaxis.

## Discussion

To our knowledge, this study is the first study to investigate the interest, intention, and preference of LAI-PrEP and their determinants among MSM in the Netherlands and in Europe. Using longitudinal data of MSM that are accessing sexual healthcare outside of a clinical trial or study context and that includes early adopters of oral PrEP prior to its formal availability, our study not only can contribute to and inform future LAI-PrEP implementation policies and actions in the Netherlands, but also settings similar to the Netherlands.

Following the DOI approach, LAI-PrEP thus does not seem complementary and would not serve a larger population as our findings showed that PrEP naïve and PrEP discontinued MSM were not more likely to intend to use LAI-PrEP compared to the early adopters and the majority oral PrEP users. These findings indicate that the introduction of LAI-PrEP may not motivate the PrEP naïve and PrEP discontinued MSM to start on LAI-PrEP as earlier research had hypothesised (21, 23). In addition, our study did not observe a repeated innovator effect from the early adopters of oral PrEP. Our analyses revealed that being an oral early adopter does not predict a higher intention for LAI-PrEP compared to the majority oral PrEP users. Also, among the current oral PrEP users, our studies did not find the innovator effect on both the intention and preference for LAI-PrEP, too. These findings may suggest that, comparing to oral PrEP as a completely novel biomedical HIV prevention tools, LAI-PrEP may be considered as just a new format/regimen of PrEP. In turn, it may not be able to motivate those oral PrEP early adopters to adopt again. Another possible reason may be positive experience with oral PrEP among MSM in the past that using oral PrEP can be sufficient to increase the quality of sex life (31). In turn, even though using LAI-PrEP can help avoiding taking medication daily/on-demand, it may not be a sufficient motivation for current oral PrEP users to switch to LAI-PrEP, regardless of their oral PrEP using status: early adopters or the majority oral PrEP users.

Yet, our study, based on 309 MSM in the Netherlands, showed that 84% of our respondents had a high/very high interest of and 63% of our respondents had a high/very high intention prior to the official introduction in the Netherlands. More importantly, 57% of our respondents still had high/very high intention to use LAI-PrEP following a 180-day daily oral PrEP obligation after discontinuing LAI-PrEP uptake, prior to the official LAI-PrEP introduction in the Netherlands. Among the included 218 current oral PrEP users, 68% of them would prefer to use LAI-PrEP. These results thus indicate an effective, available, and affordable LAI-PrEP, as the new PrEP regimen, would be welcomed in the Netherlands.

We also found that there was a very small amount of our sample living in the Netherlands who would be willing to travel to another country to obtain LAI-PrEP. This finding thus highlights the fact that the PrEP tourism observed in the beginning stage of oral PrEP (4, 5, 8, 32) may not be observed again in terms of the LAI-PrEP. One reason may be that healthcare providers, such as nurses, are needed for the LAI-PrEP injection.

In addition, we also investigated the willingness to mixing of LAI-PrEP and oral PrEP among MSM in the Netherlands. Even though this is not recommended and can raise concerns of potential drug resistance (27), we believe it can be important to investigate this scenario due to availability parameters of LAI-PrEP such as pricing of LAI-PrEP, which is assumed to be more expensive than oral PrEP. While the oral PrEP is relatively easy to access in the Netherlands, mixed use of the LAI-PrEP and oral PrEP may not be uncommon among some MSM sub-populations, such as MSM that frequently visit gay boat cruises, circuit parties or Pride events. During different periods of tourism in a year, MSM may prefer to use LAI-PrEP during the periods when they have more sex with changing partners and oral PrEP when they have less sex and partners. Our findings of a small amount of MSM were interested in using both LAI-PrEP and oral PrEP together or at different time points of a year was thus in line with this notion. Yet, due to insufficient data, we could not identify which MSM sub-populations would be more likely to mix use LAI-PrEP and oral PrEP.

However, what can be noteworthy to discuss is that our analyses revealed that the current oral PrEP users with a suboptimal adherence were more likely to prefer to use LAI-PrEP if it becomes available and affordable in the Netherlands. Even though this effect was not statistically significant, we considered this effect relevant for public health, given how the oral PrEP adherence can influence the effectiveness of oral PrEP, and especially with a very close p-value to our statistical threshold of 0.05. One reason behind this finding may be that a daily/on-demand medication taking is not required for LAI-PrEP users. Given that one of the largest barriers of a good oral PrEP adherence is the daily/on-demand medication taking (11, 18), even though our study did not find a positive association between oral PrEP suboptimal adherence and the LAI-PrEP intention among current PrEP users, oral PrEP users with suboptimal adherence can prefer to use LAI-PrEP when both PrEP regimens are available to them. This finding may thus point out the alternative targeted population for LAI-PrEP: Current oral PrEP users with suboptimal adherence. As the sub-population which using oral PrEP with the lowest oral PrEP effectiveness (16), with a higher preference for the LAI-PrEP and without the concern of the adherence issues, this population may benefit the most from using LAI-PrEP.

Our analyses also identified that MSM who were younger and ever got paid for sex were more likely to intend to use LAI-PrEP in the Netherlands. These findings were in line with the previous investigation for oral PrEP intention that a younger MSM (33), and MSM who were involved in transactional sex (4) were more likely to intend to use oral PrEP. Also, the fact that no personality trait, especially for openness and agreeableness, which were considered being associated with the innovators’ effect in oral PrEP use (24, 33, 34), was associated with both intention and preference of LAI-PrEP among both MSM in general and those who were currently using oral PrEP needs to be highlighted. These findings may indicate a similar perception of how the LAI-PrEP and oral PrEP help can prevent HIV among MSM, that the factors were associate with a higher oral PrEP intention also predict for the LAI-PrEP intention.

## Strengths and Limitations

The major strength of this study was the employment of the diffusion of innovation (DOI) approach to investigate the LAI-PrEP intention and preference among MSM and MSM who were using oral PrEP for the first time. With the DOI approach, our study was able to identify the potential target population, current oral PrEP using MSM with suboptimal adherence, for LAI-PrEP delivery when the LAI-PrEP become available in Europe. We were also able to reveal which factors determine a higher intention and preference for the LAI-PrEP, too. We thus expect our results remain its relevance to the future LAI-PrEP implementation policies and eligibility criteria policies in the Netherlands, but also other settings with similar HIV context as the Netherlands. Another strength in this study can be the cross-sectional study design nested in a cohort study. Given the known and validated PrEP use status since 2017, our data on the PrEP use status was thus devoid of information bias and was reliable for the estimation following the DOI approach.

There are few limitations that are applicable to our study. Firstly, we suggested positive experience with oral PrEP as the potential barrier for LAI-PrEP intention. However, due to insufficient data, future studies are warranted and should further investigate this with both more qualitative and empirical evidence. Secondly, we did not investigate the potential adherence to LAI-PrEP among MSM who were intending to use LAI-PrEP, due to the lack of the real-world experience with LAI-PrEP. The adherence to LAI-PrEP may be the key to the effectiveness and the potential drug resistance issues (27). If those who with a suboptimal oral PrEP adherence would also have a suboptimal LAI-PrEP adherence, our suggestion of them being the target LAI-PrEP target population may be misleading. However, as measuring LAI-PrEP adherence fell outside the scope of this research, we cannot be sure if this is the case. An updated assessment at a later stage could be warranted. Thirdly, given that the cost of LAI-PrEP remains unknown in Europe, the findings from our study were based on the scenario of availability. In terms of a much higher cost of LAI-PrEP, our finding may have resulted in an overestimation, given the established impact of the cost to oral PrEP intention in the Netherlands (25). Therefore, future studies should also assess the intention of LAI-PrEP in different scenarios of the cost of the LAI-PrEP to further investigate how the cost of LAI-PrEP may influence the intention and preference to use LAI-PrEP among MSM. Forth, given the relatively small sample size in this study, we considered a p<0.10 as relevant for our key variables of interest. Our relevant findings may be biased given the limited power. Future study should therefore investigate these topics with a larger sample size to provide more concise and more comprehensive estimations.

In summary, we observed a high interest and intention to use LAI-PrEP among MSM and a high preference for LAI-PrEP among the current oral PrEP users in the Netherlands. Early-adopters of past oral PrEP use did not show increased intention to use LAI-PrEP and neither did PrEP-naïve nor PrEP-discontinued MSM. Suboptimal oral PrEP adherence determines LAI-PrEP preference but did not determine LAI-PrEP intention. To reach the full potential of LAI-PrEP in terms of HIV prevention, a targeted LAI-PrEP implementation strategy to the current oral PrEP users with suboptimal adherence seems needed when the LAI-PrEP become available in the Netherlands.

## Data Availability

Data are available upon request.

## References

1. van Sighem A.I. Wf, Boyd A., Smit C., Matser A., Reiss P. Monitoring Report 2021. Human Immunodeficiency Virus (HIV) Infection in the Netherlands. Amsterdam: Stichting HIV Monitoring, 2021. 2021 [Available from: https://www.hiv-monitoring.nl/en/resources/monitoring-reports.

2. GGD-Amsterdam. STI, HIV and sense - PrEP 2019 [Available from: https://www.ggd.amsterdam.nl/english/sti-hiv-sense/prep/#:~:text=Dutch%20National%20programme%202019,are%20at%20risk%20for%20HIV.

3. Bierman W, Hoornenborg E, Nellen J. Nederlandse multidisciplinaire richtlijn Pre-expositie profylaxe (PrEP) ter preventie van hiv (update 2022) 2022 [Available from: https://www.soaaids.nl/files/2022-07/20220711-PrEP-richtlijn-Nederland-versie-3-update-2022.pdf.

4. Wang H, Shobowale O, den Daas C, Op de Coul E, Bakker B, Radyowijati A, et al. Determinants of PrEP Uptake, Intention and Awareness in the Netherlands: A Socio-Spatial Analysis. International Journal of Environmental Research and Public Health. 2022;19(14).

5. Hoornenborg E, Krakower DS, Prins M, Mayer KH. Pre-exposure prophylaxis for MSM and transgender persons in early adopting countries. AIDS (London, England). 2017;31(16):2179–91.

6. Coyer L, van Bilsen W, Bil J, Davidovich U, Hoornenborg E, Prins M, et al. Pre-exposure prophylaxis among men who have sex with men in the Amsterdam Cohort Studies: Use, eligibility, and intention to use. PLOS ONE. 2018;13(10):e0205663.

7. van Dijk M, de Wit JBF, Kamps R, Guadamuz TE, Martinez JE, Jonas KJ. Socio-Sexual Experiences and Access to Healthcare Among Informal PrEP Users in the Netherlands. AIDS Behav. 2021;25(4):1236–46.

8. van Dijk M, Duken SB, Delabre RM, Stranz R, Schlegel V, Rojas Castro D, et al. PrEP Interest Among Men Who Have Sex with Men in the Netherlands: Covariates and Differences Across Samples. Archives of Sexual Behavior. 2020;49(6):2155–64.

9. Krist LC, Zimmermann HML, van Dijk M, Stutterheim SE, Jonas KJ. PrEP Use in Times of COVID-19 in the Netherlands: Men Who Have Sex With Men (MSM) on PrEP Test Less for HIV and Renal Functioning During a COVID-19 Related Lockdown. AIDS Behav. 2022.

10. Jongen VW, Zimmermann HML, Boyd A, Hoornenborg E, van den Elshout MAM, Davidovich U, et al. Transient Changes in Preexposure Prophylaxis Use and Daily Sexual Behavior After the Implementation of COVID-19 Restrictions Among Men Who Have Sex With Men. J Acquir Immune Defic Syndr. 2021;87(5):1111–8.

11. Jongen VW, Hoornenborg E, van den Elshout MAM, Boyd A, Zimmermann HML, Coyer L, et al. Adherence to event-driven HIV PrEP among men who have sex with men in Amsterdam, the Netherlands: analysis based on online diary data, 3-monthly questionnaires and intracellular TFV-DP. Journal of the International AIDS Society. 2021;24(5):e25708.

12. Jonas KJ, Yaemim N. HIV prevention after discontinuing pre-exposure prophylaxis: conclusions from a case study. Frontiers in public health. 2018;6:137.

13. Coyer L, van den Elshout MAM, Achterbergh RCA, Matser A, Schim van der Loeff MF, Davidovich U, et al. Understanding pre-exposure prophylaxis (PrEP) regimen use: Switching and discontinuing daily and event-driven PrEP among men who have sex with men. EClinicalMedicine. 2020;29-30:100650.

14. Molina J-M, Capitant C, Spire B, Pialoux G, Cotte L, Charreau I, et al. On-Demand Preexposure Prophylaxis in Men at High Risk for HIV-1 Infection. New England Journal of Medicine. 2015;373(23):2237–46.

15. Dimitrov DT, Mâsse BR, Donnell D. PrEP Adherence Patterns Strongly Affect Individual HIV Risk and Observed Efficacy in Randomized Clinical Trials. J Acquir Immune Defic Syndr. 2016;72(4):444–51.

16. Jourdain H, de Gage SB, Desplas D, Dray-Spira R. Real-world effectiveness of pre-exposure prophylaxis in men at high risk of HIV infection in France: a nested case-control study. Lancet Public Health. 2022;7(6):e529–e36.

17. Whiteley L, Craker L, Sun S, Tarantino N, Hershkowitz D, Moskowitz J, et al. Factors associated with PrEP adherence among MSM living in Jackson, Mississippi. J HIV AIDS Soc Serv. 2021;20(3):246–61.

18. Sidebottom D, Ekström AM, Strömdahl S. A systematic review of adherence to oral pre-exposure prophylaxis for HIV - how can we improve uptake and adherence? BMC Infect Dis. 2018;18(1):581.

19. Zimmermann HML, Jongen VW, Boyd A, Hoornenborg E, Prins M, de Vries Hjc, et al. Decision-making regarding condom use among daily and event-driven users of preexposure prophylaxis in the Netherlands. Aids. 2020;34(15):2295–304.

20. Landovitz RJ, Donnell D, Clement ME, Hanscom B, Cottle L, Coelho L, et al. Cabotegravir for HIV Prevention in Cisgender Men and Transgender Women. N Engl J Med. 2021;385(7):595–608.

21. UNAIDS. UNAIDS responds to EU approval of a long acting HIV treatment option: “To end AIDS, share technology.” 2022 [Available from: https://www.unaids.org/en/taxonomy/term/874.

22. ViiV. EUROPEAN MEDICINES AGENCY VALIDATES ViiV HEALTHCARE’S MARKETING AUTHORISATION APPLICATION FOR CABOTEGRAVIR LONG-ACTING INJECTABLE FOR HIV PREVENTION 2022 [Available from: https://viivhealthcare.com/hiv-news-and-media/news/press-releases/2022/october/european-medicines-agency-validates-viiv-healthcare/.

23. Meyers K, Wu Y, Golub S. To switch or not to switch: anticipating choices in biomedical HIV prevention-from Oral to Long Acting Injectable PrEP-90 Men using PrEP Surveyed. 2016.

24. Rogers EM, Singhal A, Quinlan MM. Diffusion of innovations. An integrated approach to communication theory and research: Routledge; 2014. p. 432–48.

25. van Dijk M, de Wit Jbf, Guadamuz TE, Martinez JE, Jonas KJ. Slow Uptake of PrEP: Behavioral Predictors and the Influence of Price on PrEP Uptake Among MSM with a High Interest in PrEP. AIDS Behav. 2021;25(8):2382–90.

26. Landovitz RJ, Li S, Eron JJ, Jr., Grinsztejn B, Dawood H, Liu AY, et al. Tail-phase safety, tolerability, and pharmacokinetics of long-acting injectable cabotegravir in HIV-uninfected adults: a secondary analysis of the HPTN 077 trial. Lancet HIV. 2020;7(7):e472–e81.

27. Parikh UM, Koss CA, Mellors JW. Long-Acting Injectable Cabotegravir for HIV Prevention: What Do We Know and Need to Know about the Risks and Consequences of Cabotegravir Resistance? Current HIV/AIDS Reports. 2022;19(5):384–93.

28. CBS. Bevolking 15 tot 75 jaar; opleidingsniveau, wijken en buurten, 2019. 2020.

29. Rammstedt B, John OP. Measuring personality in one minute or less: A 10-item short version of the Big Five Inventory in English and German. Journal of research in Personality. 2007;41(1):203–12.

30. Lwanga SK, Lemeshow S, World Health O. Sample size determination in health studies : a practical manual / S. K. Lwanga and S. Lemeshow. Geneva: World Health Organization; 1991.

31. van Dijk M dWJ, Guadamuz T, Martinez JE, Jonas K. Quality of sex life and perceived sexual pleasure of PrEP users in the Netherlands 2020. 2020.

32. Wang Z, Fang Y, Yaemim N, Jonas KJ, Chidgey A, Ip M, et al. Factors Predicting Uptake of Sexually Transmitted Infections Testing among Men Who Have Sex with Men Who Are “Pre-Exposure Prophylaxis Tourists”-An Observational Prospective Cohort Study. Int J Environ Res Public Health. 2021;18(7).

33. Holt M, Lea T, Bear B, Halliday D, Ellard J, Murphy D, et al. Trends in Attitudes to and the Use of HIV Pre-exposure Prophylaxis by Australian Gay and Bisexual Men, 2011–2017: Implications for Further Implementation from a Diffusion of Innovations Perspective. AIDS Behav. 2019;23(7):1939–50.

34. Wu Y, Yang G, Meyers K. Acceptability, Appropriateness, and Preliminary Effects of the PrEP Diffusion Training for Lay HIV Workers: Increased PrEP Knowledge, Decreased Stigma, and Diffusion of Innovation. AIDS Behav. 2021;25(10):3413–24.

